# Four waves of cancer-related risk behaviour and personal network data from a rural Romanian community

**DOI:** 10.64898/2026.11.13.26368007

**Authors:** Marian-Gabriel Hâncean, Marius Geantă, José Luis Molina, Jürgen Lerner, Maria-Cristina Ghiţă, Bianca-Elena Mihăilă, Bogdan-Adrian Vidraşcu, Cosmina Cioroboiu

## Abstract

4P-CAN is an EU-funded project that collected longitudinal personal network data to investigate how cancer-related risk behaviours are socially organised within a rural community. Respondents (egos) describe the people they know (alters), their relationships with them, and relationships among alters. We share four waves of data collected between September 2023 and September 2025 in a Romanian rural community through interviewer-assisted network interviews and link-tracing recruitment. Five linked data tables contain 353 interviews with 142 adults, 7,602 alter roster entries identifying 2,480 people, 41,354 records of ego-reported acquaintance between alters, and recruitment-node and referral records. Linking individuals across networks and waves supports investigation of how cancer-related risk behaviours are distributed and clustered across community relationships. It also supports research on network composition, overlapping networks, reported health attributes and longitudinal measurement. Reuse should account for non-probability recruitment, dependence among observations and changes in measurement across waves.

## Background and Summary

To examine how cancer-related risk behaviours are socially organised, we need information about both individuals’ behaviours and their relationships. For example, researchers can examine whether respondents who smoke also report smoking among the people they know, and whether this pattern extends across overlapping personal networks. Repeated measurements can help trace how behaviours and relationships change together over time. Despite their value, longitudinal network datasets remain scarce, particularly in rural settings. Studies in rural Appalachia and Romania illustrate the value of examining smoking within personal networks, but both used cross-sectional designs^**1,2**^. A review of rural network case studies in the Global South similarly found a predominance of single-time-point “network snapshots” and highlighted the substantial resources required to maintain longitudinal panels^**3**^. Some longitudinal studies compare rural and urban populations. Yet their focus is on broad measures such as network size and social support, rather than repeatedly recording alter rosters, alter behaviours and relationships among alters^**4**^.

Within the 4P-CAN project on primary cancer prevention (https://4p-can.eu/), we therefore collected respondents’ reports of their own health and lifestyle behaviours, their personal relationships, and their social contacts’ attributes. Personal networks place individual attributes in a relational context: respondents (“egos”) describe the people they know (“alters”), their relationships with those people (“ego-alter ties”), and relationships among them (“alter-alter ties”)^**5**^. This design allows researchers to examine reported behaviours in relation to network composition (who surrounds an ego) and structure (how alters interact). Respondents’ self-reports remain distinct from their assessments of alters’ behaviours.

Interview-based data collection was suited to the study’s focus on personal relationships in a rural community. Participation did not require respondents to own smartphones, use online platforms or generate digital interaction records. Interviews elicited relationships and assessments that participants could report directly, including information about people who did not themselves participate. Where several respondents nominated the same person (as an alter), the resulting records connected their personal networks. Individuals could also appear both as interviewed respondents (egos) and as alters described by others. These overlapping roles create dependencies across observations and support cross-classified representations of the data^**6**^.

Other datasets in the field illustrate complementary approaches to observing social relationships. The Copenhagen Networks Study provides smartphone-derived proximity, telephone and text-message records alongside Facebook friendship data for a university population^**7**^. The DyLNet dataset combines repeated sensor-based proximity observations in a French preschool with sociodemographic and language-development information^**8**^. These datasets differ from ours in their populations and recording methods and capture complementary aspects of social relationships.

We share four waves of data collected between September 2023 and September 2025 in a Romanian rural community. Respondents were recruited through link tracing from six purposively selected seeds. Five linked data tables contain respondent interviews and attributes, alter attributes and ego-alter relationships, ego-reported acquaintance between alters (alter-alter ties), recruitment-node attributes and directed recruitment referrals. Person identifiers connect observations across tables and waves, while respondent and wave identifiers preserve the source and occasion of each report.

According to our research design, repeated data collection provides observations of study participation, alter nominations and reported attributes over time^**9**^. However, the elicitation procedures changed across waves^**10**^. Waves 1,3 and 4 used a fixed nomination target, while wave 2 elicited alters without an upper limit. Wave 4 distinguished returning respondents who reviewed historical rosters (their previously elicited lists of alters) from first-time respondents who generated new rosters. Documenting these differences helps users distinguish changes in reported networks from changes in how they were elicited.

Earlier studies used selected parts of this dataset to examine methodological questions and health-related behaviours. A participatory methods article describes the preparation and adaptation of data collection for the first three waves^**9**^, and a separate methodological study compares namegenerator designs in the first two waves^**10**^. Substantive applications examine smoking^**2**^, processed-food intake^**11**^, and online media use and vaccination^**12**^. Another publication describes returning findings through a community art gallery^**13**^. Earlier data deposits contain study-specific subsets or derived outputs associated with previous publications^**14-19**^. These deposits should not be assumed to contain the same observations, identifiers, variables or coding conventions as the consolidated four-wave dataset described here. The present Data Descriptor documents the consolidated four-wave data release^**20**^ and the collection, linkage, curation and validation procedures.

Potential reuse includes examining how reported cancer-related risk behaviours are distributed and clustered within personal networks and across networks linked by shared alters. Within the dataset, individuals appearing as both egos and alters enable comparisons between self-reports and reports provided by others. The data also support research on overlapping networks, repeated nominations, recruitment patterns, and the sensitivity of network descriptions to eligibility criteria and elicitation procedures. Such analyses can inform questions relevant to community-based prevention, although the dataset itself does not establish intervention effectiveness. The dataset constitutes neither a probability sample of residents nor a census of community relationships. Analyses should accommodate dependence arising from repeated observations and shared individuals, distinguish reporting sources, and account for changes in measurement across waves. Analyses of social selection and influence require models of changes in both relationships and behaviours, and causal interpretations depend on assumptions about network coverage, measurement, and confounding.

## Methods

### Study design and recruitment

We recruited participants using a network-based link-tracing sampling design. We purposively selected six individuals as seeds, seeking variation in sex, age, income, education, and employment sector to encourage varied referrals. Recruitment records include wave-1 referrals from all six seeds. Serving as a seed did not require completing a personal-network interview: five seeds completed at least one interview during the study, whereas one contributed a referral without being interviewed. Eligibility for interview participation required being aged 18 years or older, having resided in the rural community for at least six months, being able to speak and understand Romanian, and being able and willing to provide informed consent. Community engagement, including field observations and immersion between May and September 2023, preceded recruitment and helped the research team establish connections with residents.

Participants were asked to recommend and help recruit other eligible residents. Initially, the team requested at least five referrals (waves 1,2), but this became an approximate target (waves 3,4) as successive interviews showed that respondents often found it difficult to recommend that many people. The research team contacted recommended individuals and arranged interviews according to their availability and the team’s fieldwork capacity. The recruitment procedure allowed up to three invitation attempts per person in each wave (the data deposit does not document the actual number of attempts for every nominee). In wave 4, returning respondents also provided referrals, while first-time respondents followed the wave-3 referral procedure.

The study used an open-cohort design, combining follow-up interviews with the inclusion of first-time respondents across four waves. Respondents who missed a wave could participate again in a later wave. Follow-up prioritised previously interviewed respondents. When they were unavailable, the team sought interviews with available individuals from the recruitment pool. Scheduling and fieldwork constraints prevented the team from interviewing some recommended individuals. Recorded referral outcomes do not provide a complete history of contact attempts or reasons for nonparticipation among previously interviewed respondents.

Data collection took place on 12-23 September 2023 (wave 1), 21 March-18 April 2024 (wave 2), 20 March-16 April 2025 (wave 3), and 14-21 September 2025 (wave 4). The main field visits for waves 2 and 3 ended on 1 April 2024 and 5 April 2025, respectively. In each of these waves, one rescheduled interview was completed after the main field visit. Participants received no financial compensation.

The study was reviewed and approved by the Ethics Committee of the Center for Innovation in Medicine (INOMED; decisions D001/09-06-2023, D001/19-01-2024 and D002/03-02-2025). The 2025 decision covered both the March-April and September 2025 fieldwork periods. Participation was voluntary. Written informed consent was obtained from all participants for study participation and for the use and sharing of data for scientific research, including future research.

### Interview administration

All interviews, including rescheduled ones, were conducted one-to-one, face-to-face, and in Romanian. Each interviewer used a laptop running Network Canvas to record responses during the interview. Versions 6.0.0 and 6.5.3 were used in wave 1 and waves 2-4, respectively^**9,21**^. Interviewers attended specialised Network Canvas training sessions delivered by the software’s developers at the International “Winter Course” titled “Personal Networks: Theory, Methods and Applications in Barcelona” (Universitat Autònoma de Barcelona; https://webs.uab.cat/egolab/12th-summer-course-personal-networks/). Before each fieldwork wave, the team pretested the protocol in-house for duration and question interpretation. During fieldwork, the team discussed unanticipated situations and coordinated administration. Interviews within a wave followed the same protocol, including its routing (skip logic) for different respondent groups.

The research team sought, where feasible, to assign returning respondents to the team member who had interviewed them previously. To assess interviewer continuity, we compared the interviewer recorded for each return interview with the interviewer who had conducted that respondent’s most recent prior interview. Under this definition, the same team member conducted the current and most recent prior interviews for 33 of 68 returning respondents in wave 2 (48.5%), 25 of 76 in wave 3 (32.9%), and 16 of 67 in wave 4 (23.9%). The denominators include respondents returning after a missed wave.

For returning respondents, previously recorded identifying information, including reported date of birth and recorded sex, was carried forward. Access to earlier health and lifestyle responses was restricted: the team member conducting the current interview could not view those previous answers. Health and lifestyle questions followed the routing specified in each wave’s questionnaire. In wave 4, questions on sun protection, alcohol consumption and primary prevention were restricted to first-time respondents. The number of cigarettes smoked per day was asked only of first-time respondents who reported current regular or occasional smoking. Within the primary-prevention section, questions about screening in the previous 12 months and the type of cancer screened for were asked only of respondents who reported screening in the previous five years. The question on perceived future cancer risk was not asked of respondents known to be cancer patients.

### Personal network elicitation

Table 1 summarises the written questionnaire instructions in each wave for naming social contacts (alters) and, where applicable, reviewing contacts named in previous waves.

**Table 1.**
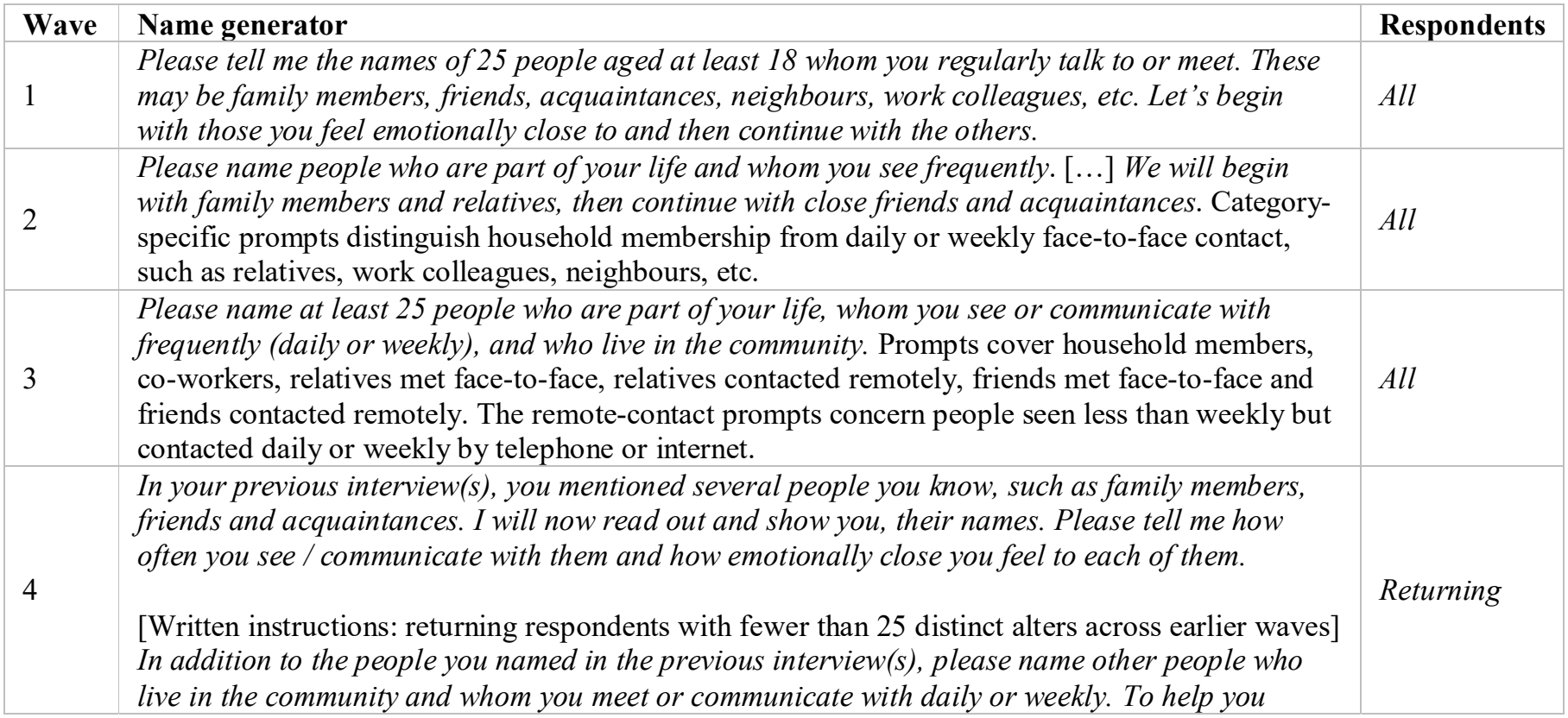
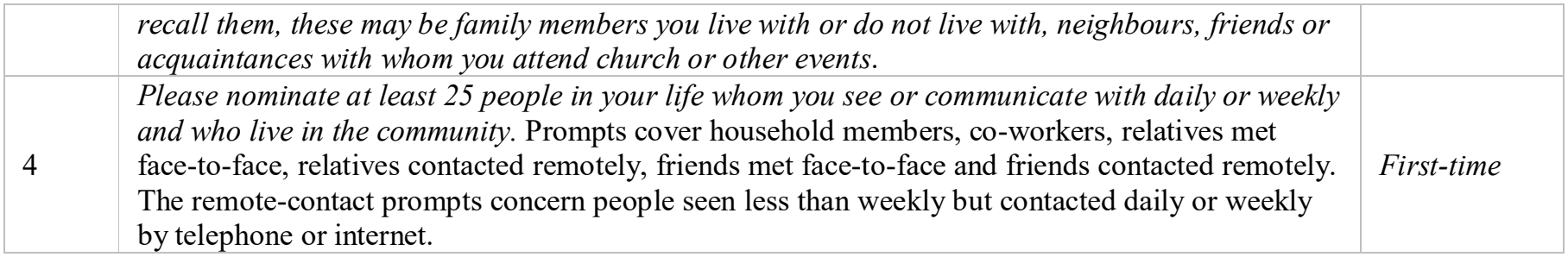
English summaries of the instructions for naming social contacts and reviewing previously named contacts across the four waves. The full questionnaires specify which contacts could be nominated in each relationship category and when particular questions were asked or skipped.

In wave 1, respondents were asked to name people with whom they regularly talked or met and to list them in order of emotional closeness. The target was 25 names, but respondents with shorter lists remained in the study. The curated wave-1 data contain 1,967 alter nominations from 83 respondents, with a median of 25 alters per respondent and a range of 10-25. Of the 83 rosters, 61 contain 25 names, and 22 contain fewer than 25. The nomination instructions did not explicitly require these contacts to live in the rural community.

In wave 2, respondents were guided through a series of prompts to name contacts across relationship categories, including household members, relatives, neighbours and co-workers. For co-workers, relatives outside the household, friends and other contacts, the prompts specified daily or weekly face-to-face meetings. The household prompt referred to living together and meeting face-toface, without specifying how often those meetings should occur. The research design assumed that living in the same household inherently implies daily interaction. This prompt also required respondents to list all household members. Alter entries under 18 were not included in the alter roster. Returning respondents created new lists without seeing their nominations from the previous wave. There was no upper limit on nominations, and interviewers did not explicitly encourage respondents to reach 25 names. The curated data contain 1,487 alter entries from 94 respondents, with a median of 16 alters per respondent and a range of 4-31. Six rosters contain more than 25 names. As in wave 1, the prompts did not explicitly require all nominated contacts to live in the community.

In wave 3, returning respondents again named their social contacts without seeing their earlier lists. The nomination instructions explicitly requested contacts living in the community and distinguished between face-to-face meetings and remote communication. Separate prompts asked about household members, co-workers, relatives and friends. Face-to-face prompts specified daily or weekly meetings, while remote-contact prompts asked about relatives and friends seen less than weekly but contacted daily or weekly by telephone or internet. Although the written introduction requested “at least 25” names, some of the respondents did not reach the target. The curated data contain 1,623 alter entries from 89 respondents, with a median of 17 alters per respondent and a range of 2-48. Twenty-eight rosters contain more than 25 names.

Wave 4 used separate procedures for returning and first-time respondents. Returning respondents were shown the names of previously nominated alters, which interviewers also read aloud, and asked to review each relationship. They reported their current frequency of contact, emotional closeness and each alter’s current place of residence. Contact-frequency options were daily or weekly, monthly, less than monthly, and no contact. When contact had ceased, respondents could give reasons such as death, divorce, moving away, estrangement or fewer opportunities to meet. The 67 returning respondents contributed 2,218 retained roster entries, with a median of 34 alters per respondent and a range of 4-63. These entries include 63 reports of no current contact and 703 reports of alters living outside the community. Retained entries are nominations included after data checking and curation.

The written wave-4 instructions targeted supplementary nominations from returning respondents with fewer than 25 distinct alters across earlier waves, aiming to bring the roster to 25. In field administration, all returning respondents were invited to name additional active contacts, with more prompting for those below that threshold. The supplementary prompt asked for community residents whom respondents met or contacted daily or weekly, using family, neighbours, friends and acquaintances as recall aids. Fourteen returning respondents contributed 74 entries explicitly marked as supplementary nominations. All 74 were recorded as daily or weekly contacts: 73 were recorded as living in the community and one as living elsewhere. Returning respondents were asked to estimate how many years they had known each alter, including both previously nominated contacts and supplementary nominations. Among the supplementary nominations, the median reported duration of acquaintance was 24.5 years. These durations describe how long contacts had been known, not when regular contact began or why they had not been nominated earlier. The 20 first-time respondents in wave 4 received the introduction and nomination prompts used in wave 3. They contributed 307 alter entries, with a median of 14 alters per respondent and a range of 6-25.

Together, these procedures changed recall assistance, contact and residence criteria, and the presentation of earlier nominations. Their implications for longitudinal comparisons are discussed in **Usage Notes**. Additionally, we underline that personal-network nominations and recruitment referrals denote different relationships: naming someone as an alter does not necessarily imply recommending that person for study participation.

**Fig. 1** illustrates how the nomination procedures shaped the recorded data for one respondent interviewed in all four waves. This respondent’s lists of nominated contacts contained 15, 11, 10 and 25 alters in waves 1-4, respectively. In wave 2, six of the 11 alters had also been nominated in wave 1, while five were recorded for the first time. The wave-3 roster included six alters from wave 2, one nominated in wave 1 but absent from wave 2 (A01), and three recorded for the first time. In wave 4, this respondent reviewed all 23 distinct alters nominated across waves 1-3 and added two further individuals. The interviewee reported daily or weekly contact with 20 alters, monthly contact with four, and no current contact with one previously nominated alter. The respondent was also asked to estimate how many years they had known each of the 25 alters, including both previously nominated and newly added contacts. The two newly added alters had reportedly been known for 35 and 74 years, showing that newly recorded nominations can represent longstanding relationships. The alter-alter graphs contained 51, 42 and 42 ties, indicating that, according to the respondent, the two alters knew each other in waves 1-3, respectively. Relationships among alters were not reassessed for this returning respondent in wave 4.

**Fig. 1.**
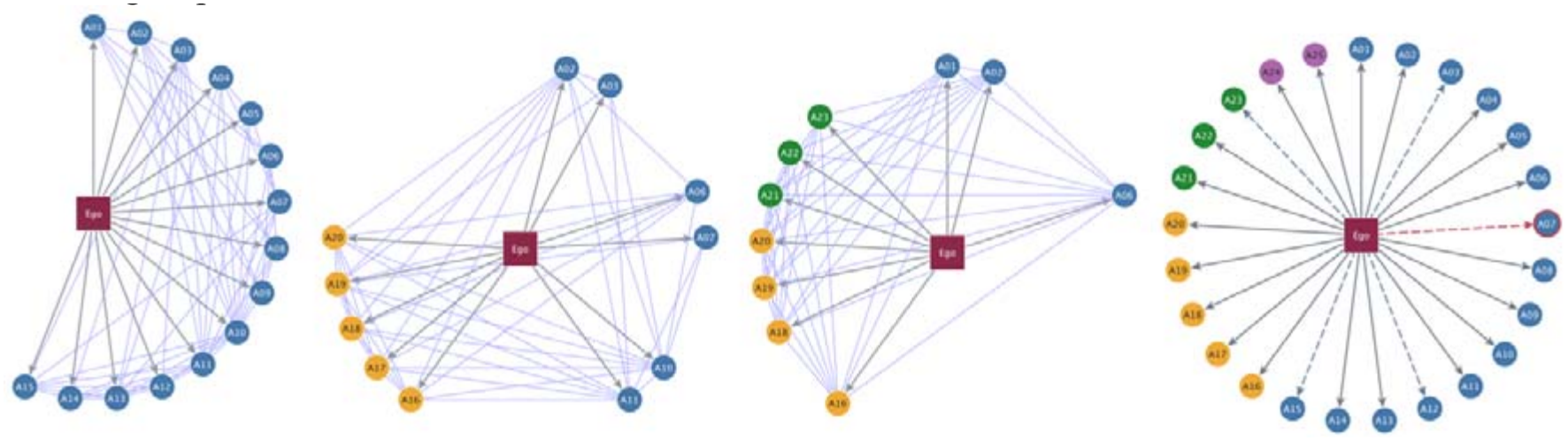
Personal-network elicitation across four waves for one illustrative respondent. Panels represent waves 1-4 from left to right. The burgundy square denotes the respondent (ego), and labelled circles denote contacts (alters). Labels identify the same individuals across panels. Node colours indicate the wave in which an alter appeared first in this respondent’s roster: blue (wave 1), orange (wave 2), green (wave 3), and purple (wave 4). Arrows from ego identify nominations in the wave-specific roster. Thin light-blue lines connect alters whom the ego reported as knowing each other in waves 1-3 (the questions describing these relationships differed across waves). In wave 4, solid grey arrows indicate daily or weekly contact, dashed blue arrows indicate monthly contact, and the dashed red arrow and red node border identify the alter with no current contact (A07). The fourth panel contains reviewed entries and additional nominations. Alter-alter ties were not reassessed for this returning respondent in wave 4. The respondent was selected for illustration, not representativeness. First appearance in a roster does not establish when a relationship began, and omission from a freshly elicited roster does not establish relationship dissolution. Node positions serve graphical readability and do not measure geographical distance or emotional closeness. The wave-4 panel does not represent disconnected alters, as relationships were not reassessed and previous alter-alter ties were ignored.

### Respondent and alter attributes

Table 2 summarises the respondent, alter and relational measures retained in the data tables and their coverage across the four waves. The accompanying questionnaires provide the full wording of the questions, response categories and routing. In wave 3, each respondent evaluated all eight food categories separately. The data release contains one frequency field for each category.

**Table 2.** Respondent, alter and relational measures retained in the released tables and their wave coverage. W1-W4 denote waves 1-4; W4N denotes first-time respondents, and W4R denotes returning respondents. Coverage refers to retained fields and questionnaire eligibility, not universal administration, complete responses or identical wording. Exact ages are replaced by age groups; respondent Body Mass Index (BMI) is derived from self-reported height and weight. Photographic body-size responses are image selections, not calculated BMI.

| **Domain** | **Measures and wave coverage** |
|---|---|
| Respondent sociodemographic and household characteristics | Age group, recorded sex, grouped education, occupation, relationship status and personal/household income (W1-4). Household size and child counts are excluded in the released tables. Education was asked only of first-time respondents in W2-W3 and was included for both respondent groups in W4. Education was confirmed for returning participants. |
| Respondent health | Self-rated health (W2-4). Reasons for the rating and personal/family cancer and cardiovascular histories are excluded. |
| Respondent healthcare and prevention | Routine check-ups, cancer screening and usual response to a health problem (W3; W4N), with additional screening follow-up filters. Detailed W1 provider and visit information is excluded. |
| Respondent vaccination | COVID-19, HPV and hepatitis B vaccination and sources of vaccination advice (W1). |
| Respondent body size | Self-reported height and weight, derived BMI and BMI categories (W1-4); photographic self-assessment of body shape (W4). |
| Respondent food and dietary practices | Processed-food consumption and motives (W1-2); eight food-group frequency items (W3-4); salt, sugar, meat and water items (W1); retained meal narratives/timing, perceived ability of household income to cover desired food and healthy-meal suggestions (W2); dieting and perceived dietary healthiness (W2-4); role-only cooking/shopping-list responses (W2-3). W4 cooking/shopping narratives and person-code links are excluded. |
| Respondent smoking and other behaviours | Smoking status and sport (W1-4, with different sport definitions); tobacco types and smoking history (W1); first cigarette relative to the first meal, asked of current smokers (W2); cigarettes per day, asked of current regular or occasional smokers (W3; W4N). Alcohol and sun protection (W1, W3; W4N); work/home physical activity (W1); sleep/wake times (W2). |
| Respondent information sources and community activities | Health-information sources (W1, W3; W4N); preferred providers of cancer-prevention information and campaign recall/preferences (W1); perceived cancer knowledge and risk (W3; W4N, with risk eligibility filters); overall project evaluation (W3-4); retained future-event preferences, communication preferences and prospective involvement (W4). Event attendance and attendance-revealing follow-ups are excluded. |
| Alter sociodemographic attributes | Ego-reported age group, recorded sex and grouped education (W1-4); grouped relationship status (W1); grouped occupation (W2-3; W4N). |
| Ego-alter relationships | Relationship and kinship descriptions (W1); household membership (W1-3; W4N); coworker, relative and friend nomination categories (W2-3; W4N). Additional nomination categories for relatives and friends contacted daily or weekly by telephone or internet, but met face to face less than weekly (W3; W4N). |
| Ego-alter contact and roster review | Meeting frequency (W1); daily interaction (W3; W4N); emotional closeness (W1, W3-4); years known (W4). The W4R roster review records current contact frequency, residence, supplementary-nomination status and reasons for no contact. |
| Ego-alter meals and advice | Shared meals (W2-4); special food preparation for household members (W2-3; W4N); health advice (W3-4); attempts to persuade the respondent to stop smoking or change diet (W3; W4N), with the smoking item restricted to current or former smokers. |
| Alter health and behaviours | Ego-reported smoking, sport, dieting, processed-food consumption and photographic body shape (W1-4); alcohol and vaccination (W1); health rating (W3-4). The W4R health/lifestyle block was intended for daily/weekly contacts. Reported cancer and heart-disease diagnoses are excluded. |
| Alter-alter assessments | Only pairs reported by the ego to know each other are retained. W1 distinguishes acquaintances, regular friends and close friends. W2 distinguishes interaction less than weekly from interaction at least weekly. W3 and W4N distinguish daily/weekly, monthly and less-than-monthly interaction, and acquaintance with unknown interaction frequency. W4N additionally distinguishes acquaintance without interaction. No updated assessments were collected for W4R. |

Respondents reported their own health and behavioural information (medical documents were not requested). Height and weight were self-reported in all four waves. Direct measurement using scales and a height measure was initially planned for wave 1, but the team considered this too intrusive during fieldwork and used self-reports instead. Respondent age was calculated from the reported date of birth, while alter age was reported by the respondent who nominated that person. Both respondent and alter ages are provided as age groups in the released dataset. Respondent body mass index (BMI) was calculated as weight in kilograms divided by height in metres squared and rounded to one decimal place.

When an alter also participated as a respondent, information supplied about that person by others was stored separately from their own responses. Alter sex was recorded according to the questionnaire’s interviewer instructions, using names where informative and seeking clarification when needed. The wave-1 respondent sport question included walking and cycling as possible activities, whereas the wave-2 to wave-4 questions asked about daily or weekly sport and explicitly excluded walking, strolling, fishing and chess.

In all four waves, respondents assessed alters’ body size using sex-specific photographic showcards^**22**^ with ten images labelled A-J. They pointed to the image they considered most representative of each alter, without seeing BMI values or weight-category labels. In the data tables, the recorded codes, ranging from 1 to 10, identify the selected images and do not represent measured BMI. In wave 4, respondents also used the same showcards to assess their own body shape.

In wave 4, returning respondents were instructed to answer the alter health and lifestyle questions, together with the shared-meal question, only for contacts they met or communicated with daily or weekly. Questions in the preceding roster review, including health-advice sources and years known, were not subject to that contact-frequency restriction. First-time respondents answered a separate set of questions about their alters, including occupation, which was not asked of returning respondents in that wave. A variable’s presence in the wave-4 data therefore does not mean that the question was asked about every alter.

The released data tables include responses in their original form, recoded variables, derived measures, and identifiers linking observations across tables and waves. Recoded and derived variables include grouped sociodemographic measures, respondent BMI and BMI categories, and interview duration. The variable inventory lists the released variables, their formats and available values. The recoding dictionary documents category groupings, code-to-label correspondences and information on derived measures. The variable inventory and recoding dictionary are available in the data deposit^**20**^.

### Relations among alters

Respondents reported on relationships among the people they had nominated, shown by the light blue lines connecting alters in **Fig. 1**. These alter-alter assessments reflect the respondent’s knowledge of the relationship between each pair of contacts. The questions used to assess these relationships varied across waves. The protocol asked respondents to assess every distinct pair of nominated alters, considering each pair once without assigning a direction to the relationship.

In wave 1, respondents classified each pair of alters as “very good (close) friends”, “regular friends”, “acquaintances”, or “do not know each other / do not know or no response”. The last option combined definite non-acquaintance, uncertainty and nonresponse, so these cannot be distinguished from that response alone. Wave 2 offered three options: the alters interact “at least once a week”, “less often (than once a week)”, or “do not know each other”. Unlike waves 3 and 4, it did not provide a separate written response category for uncertainty about whether the alters knew each other. The question concerned face-to-face meetings between alters, without the respondent necessarily being present. The wave-3 and wave-4 alter-alter questions used the same face-to-face framing, even though their name generators also included remote contacts (see **Table 1**). Wave 3 distinguished six responses: the alters definitely did not know each other; the respondent was unsure whether they knew each other; they knew each other, but the respondent did not know how frequently they interacted; or they interacted daily or weekly, monthly, or less than monthly (“once every few months/a few times a year”). In wave 4, only first-time respondents answered the alter-alter questions. The questionnaire retained the six wave-3 categories and added a seventh for alters who knew each other but did not interact. Returning respondents did not provide updated alter-alter assessments in wave 4 (see **Fig. 1**).

### Tabular data structure integration and curation

After each wave, the interview exports were brought together in a database using ‘R code’ provided in the Network Canvas documentation (https://documentation.networkcanvas.com/en/analyze-data/working-with-data). The research team then cleaned the data and matched people manually across interviews, roles and waves. This work produced the curated tables used for the subsequent checks and preparation of the public release. The shared person identifier, ‘node_code’, was assigned through this manual review; it was not generated by the import procedure.

The team manually compared records using all available information, including names, ego-alter and alter-alter relationships, and sociodemographic characteristics. Records judged to refer to the same person were assigned the same identifier (’node_code’). When the information was insufficient to establish a match, separate codes were retained. Two teams of two researchers carried out the matching and review. The first pair assigned the codes, checked each other’s work and discussed uncertain cases. The second pair reviewed the difficult cases referred to them; they did not independently recode every record. Because the process involved joint discussion rather than independent coding of all records, no independent measure of agreement between coders is available. A separate log of uncertain potential matches was not maintained during the initial matching process.

The combinations needed to link records, including the separate role of ‘unique_alterID’, are specified in Data Records. Raw operational Network Canvas identifiers were removed from the data release. **Fig. 2** illustrates these links among data tables using example records. The highlighted cells trace person 2 as an interviewed respondent, an alter nominated by respondent 1 in waves 1 and 2, and an endpoint in that respondent’s alter-alter assessments. Person 1232 is described by two respondents, whose identifiers keep their reports separate. The recruitment network shows person 1 nominating persons 2 and 3, and person 2 nominating person 4. Person 3 is nominated in wave 1 and interviewed in wave 2, illustrating that referral and interview need not occur in the same wave. Alter-alter pairs are unordered: (i, j) and (j, i) represent the same relationship, despite the endpoint names “source” and “target”. Recruitment referrals are directed: a referral from i to j is distinct from one from j to i.

**Fig. 2.**
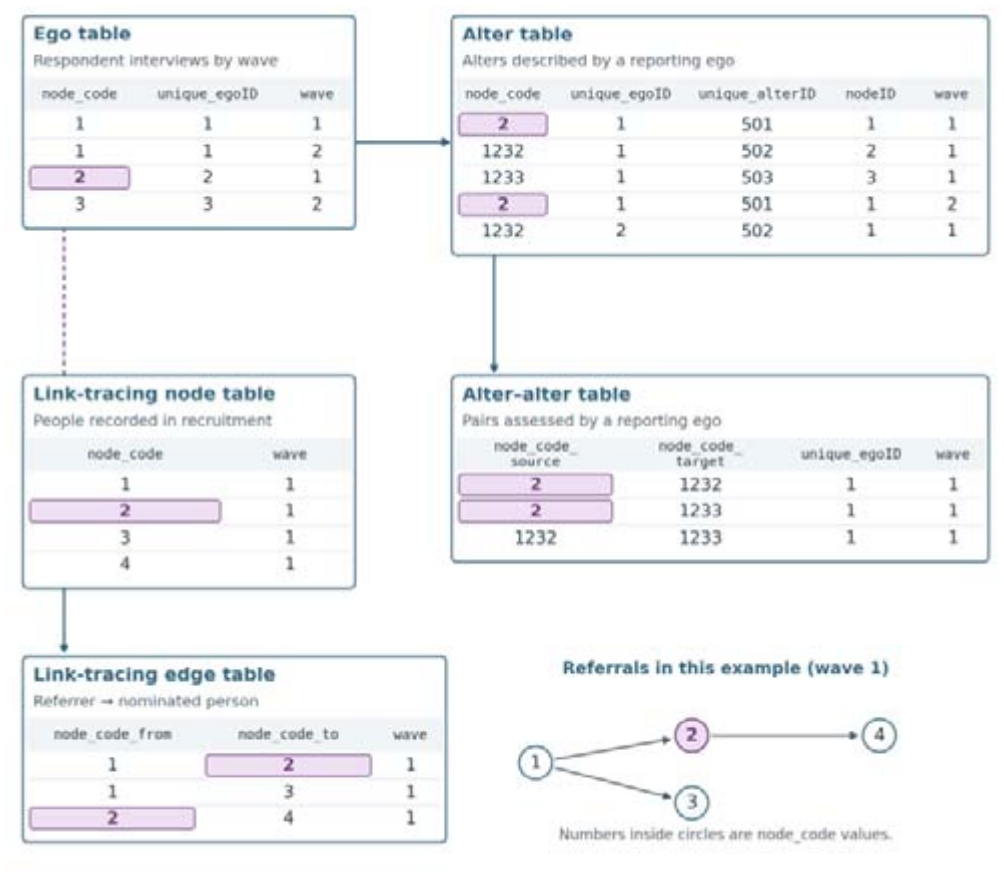
Links among the five data tables. Purple cells highlight records referring to the same person (’node_code’ = 2). Solid arrows between tables indicate the data needed to construct personal networks and the recruitment network. Dashed lines connect records referring to the same person. Each alter-alter record identifies a pair whom one respondent reported as knowing each other in one wave; the same pair may be reported by different respondents or in different waves. Recruitmentedge endpoints are linked to recruitment-node records using the person’s ‘node_code’ and ‘wave’. In the small recruitment network, arrows point from the referrer to the nominee. The displayed records are illustrative and should not be read as an extract of these individuals’ actual records in the release.

The questionnaires used ‘-98’ for “not applicable” and ‘-99’ for “do not know” or “no response”. During data preparation, these codes were replaced with missing values (’NA’) or, for some variables, explicit labels indicating uncertainty. Missing values also occur when a question was not asked in a particular wave or was not applicable to a respondent or alter. An ‘NA’ alone does not distinguish questionnaire routing, nonresponse or curation; interpretation should follow the relevant questionnaire instructions.

### Curation before release preparation

For the public release, we retained only alter-alter records indicating that, according to the respondent, the two alters knew each other. Records coded as non-acquaintance or uncertainty about whether the alters knew each other were excluded. In wave 1, the relevant response category also included uncertainty and nonresponse, so its removal cannot be interpreted as excluding only definite non-acquaintance. Interview durations above 240 minutes were treated as recording errors and set to missing; all other responses from those interviews were retained. The released alter-alter table contains 41,354 records: 14,727, 9,375, 14,917 and 2,335 in waves 1-4, respectively. Records indicating that alters knew each other but that their interaction frequency was unknown remain in waves 3 and 4. Wave 4 also retains records indicating that alters knew each other but did not interact.

### Preparation of the released tables

We prepared versions of the five curated tables for release with identifying and sensitive information removed. We deleted personal names, telephone numbers, dates of birth, exact interview and export timestamps, raw operational identifiers and identity fields. We also excluded household size and numbers of children; personal and family medical histories; reported cancer and cardiovascular diagnoses among alters; detailed information about healthcare providers and visits; community-event attendance and related follow-up questions; and selected free-text response fields containing names, identifying contextual information or personal health details. In three otherwise retained free-text variables, one response per variable was replaced with a missing value because it contained personal health information. One response was removed from each of ‘ego_food_breakfast_w2’, ‘ego_cancer_screening_type_w3’ and ‘ego_events_want_w4’. No respondent records were removed. The questionnaires document the broader information originally collected.

Respondent ages and alter ages reported by respondents were grouped into 18-35, 36-55 and 56+ years. Education was harmonised into three categories: up to eight years of schooling, nine years to post-secondary non-tertiary education, and university education. The questionnaires for waves 2-4 already used three education categories. Occupation was grouped as employed/self-employed, retired or other reported status. Personal and household income were grouped as,500 RON, >2,500 to 6,000 RON, and >6,000 RON. Relationship status distinguished those in a relationship from those not in a relationship, with a separate category for unclear original responses where applicable. Each alter report was recoded separately, preserving differences between respondents’ descriptions of the same person. Each interviewer received a pseudonymous code that remained consistent across waves.

Preparing the tables for release did not change the number of rows in the curated input tables. Person codes, wave assignments, reporting-respondent identifiers and network endpoints were also preserved. This step reduced identifying and sensitive information and was separate from the earlier curation of alter-alter records and correction of invalid interview durations. In summary, interviewers recorded responses on laptops using Network Canvas. The interview exports were then combined using R code provided in the Network Canvas documentation. The research team manually cleaned the data and matched records referring to the same person across interviews and waves. The resulting tables were subsequently prepared for public release by removing or grouping identifying and sensitive information, followed by checks of the released files.

## Data Records

The dataset is deposited in Zenodo^**20**^ as five linked data tables, each provided in RDS and CSV formats. **Table 3** identifies each file’s dimensions and observational unit. Because the tables describe different types of observations, their row counts do not represent equivalent numbers of distinct individuals

**Table 3.** Released data files, their numbers of rows and columns, and the type of observation represented by each row. RDS filenames are shown; the corresponding CSV files use the same names with the ‘.csv’ extension.

| File | Rows × columns | Observational unit |
|---|---|---|
| ego_table.rds | 353 × 370 | One respondent interview |
| alter_table.rds | 7,602 × 162 | One alter in one respondent’s wave-specific roster |
| alter_alter_table.rds | 41,354 × 12 | One pair of alters reported to know each other by one respondent in one wave |
| linktracing_node_table.rds | 383 × 27 | One person recorded in recruitment in one wave |
| linktracing_edge_table.rds | 338 × 3 | One directed recruitment referral in one wave |

Each record is identified by a combination of variables. In the ego table, ‘unique_egoID’ and ‘wave’ identify a respondent’s interview. In the alter table, adding the nominated person’s ‘node_code’ identifies an alter entry within that interview; ‘nodeID’ can be used instead of ‘node_code’ for this purpose. Each released alter-alter record is identified by ‘unique_egoID’, ‘wave’ and the unordered pair of ‘node_code_source’ and ‘node_code_target’. Each endpoint links to the corresponding alter entry using the same reporting respondent, wave and person code. Recruitment-node records are identified by ‘node_code’ and ‘wave’, while recruitment-edge records are identified by ‘node_code_from’, ‘node_code_to’ and ‘wave’. The separately assigned ‘unique_alterID’ should not be substituted for these combinations.

Each table includes a ‘wave’ variable, and many substantive variables have names ending in ‘_w1’, ‘_w2’, ‘_w3’ or ‘_w4’ to indicate the wave they describe. The values of these variables are missing in rows belonging to other waves. Many ungrouped items are provided as both numeric response codes and labelled categories. In the RDS files, grouped demographic variables are stored as factors, with ordered categories for age, education and income. For categorical variables stored as R factors, use the category labels rather than the internal numbers used by R to store them. The corresponding CSV columns contain these labels (separate numeric response-code columns retain their recorded codes).

The accompanying README explains how the files are organised and how to read them in R. The variable inventory (’variable_inventory.csv’) lists variable names, formats, category labels, recorded-value counts and wave coverage. The variable-change record (’variable_changes.csv’) documents excluded and replaced fields, while the recoding dictionary (’recoding_dictionary.csv’) documents category groupings, derived measures and export conventions. Supporting files describe table dimensions and links between tables. The wave-specific questionnaires document the wording and routing beyond the variables retained in the data release.

### Data overview

Across the four waves, 142 respondents contributed 353 interviews: 83, 94, 89 and 87, respectively. Forty-two respondents participated in one wave, 29 in two, 31 in three and 40 in all four. Among the 87 respondents interviewed in at least two of the first three waves, a mean of 6.34 alters per respondent were nominated in every wave they attended (SD = 3.68; median = 6; range = 1-22). Among the 57 respondents interviewed in all three waves, the mean was 6.19 alters (SD = 3.58; median = 6; range = 1-15). Wave 4 was excluded from this calculation because returning respondents reviewed historical rosters. These counts describe repeated nominations under the different elicitation procedures used in waves 1-3.

The recruitment table contains 338 referral records (168, 88, 62 and 20 by wave), identifying 269 distinct nominees. Pooling repeated directed pairs gives the network in **Fig. 3**, with 274 individuals and 317 arcs. Five of the six seeds nominated others but were not nominated, accounting for the five additional individuals; the sixth seed is already among the nominees. Overall, 82 individuals supplied referrals, and 45 nominees had more than one distinct referrer. Ignoring direction, the network has a main component of 272 nodes and an additional component of two individuals. Of its 274 individuals, 142 were interviewed, and 132 were never interviewed. Among the 317 arcs, 173 connect two individuals interviewed at least once, 143 run from an interviewed to a never-interviewed person, and one runs from a never-interviewed seed to an interviewed person. Participation refers to the full study, not necessarily the wave of referral. Arcs document referrals, not contact or interview completion.

**Fig. 3.**
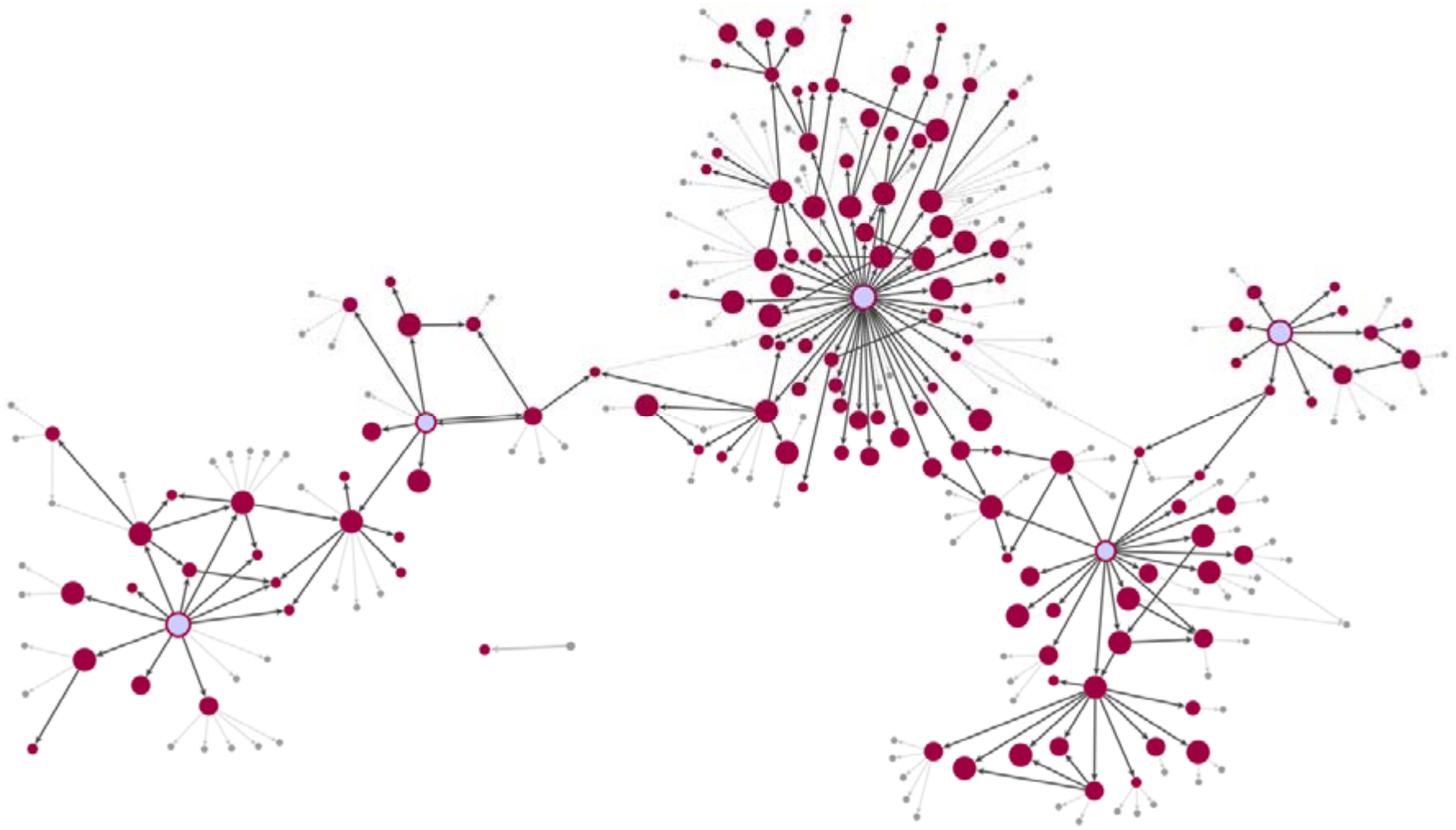
Recruitment referrals and interview participation across four waves. Nodes represent 274 individuals, and 317 directed arcs represent distinct referral pairs, with arrows pointing from the referrer to the nominated person. Node size indicates the number of waves in which an individual participated (0-4). The six seeds are light blue; the five interviewed seeds have a dark red border. Among the remaining nodes, dark red indicates individuals interviewed in at least one wave, and pale grey indicates those never interviewed. Dark grey arcs connect two individuals who were interviewed at least once. Light grey arcs connect an interviewed and a never-interviewed individual, including the referral from the never-interviewed seed. Participation is defined across all four waves and does not imply that connected individuals were interviewed in the same wave.

### Technical Validation

#### Record uniqueness and linkage

We checked the released tables for duplicate records and links that could not be matched across tables. Each respondent-wave combination occurred only once in the ego table, and no alter’s ‘node_code’ appeared more than once within the same respondent’s roster in a given wave. Alter-alter records contained no pairs linking a person to themselves and no repeated unordered pairs within an interview. Recruitment-node records were unique for each person and wave, and referral records were unique for each directed referrer-nominee pair and wave. Every reporting respondent in the alter and alter-alter tables matched an ego interview in the same wave. Both alters in every alter-alter assessment matched entries in that respondent’s roster for that wave. Every referrer and nominee in the recruitment-edge table also matched a recruitment-node record in the same wave.

The linkage checks confirm that the released tables use the documented identifiers consistently. Decisions about which records referred to the same person were made through the manual matching and team review described in **Methods**. ‘unique_alterID’ and ‘node_code’ follow separate assignment schemes, so the checks did not require their values to be equal.

#### Content and routing checks

The released tables contain no literal ‘-98’ or ‘-99’ missing-value codes. All retained alter-alter response categories indicate that, according to the ego, the two alters knew each other. None indicates non-acquaintance or uncertainty about acquaintance. Records indicating that alters knew each other but that their interaction frequency was unknown remain: 1,192 in wave 3 and 57 in wave 4. Another 179 wave-4 records indicate that the alters knew each other but did not interact.

For returning respondents in wave 4, the data contain no updated alter-alter assessments or alter occupation values, consistent with the data-collection design. Their reports of alter health and smoking are recorded only for contacts they met or communicated with daily or weekly. Questions restricted to first-time respondents, including those in the primary-prevention module, have missing values for returning respondents because they were not asked.

Interviewer codes are available for all 353 interviews, allowing checks of whether respondents were interviewed by the same interviewer across waves, as reported in **Methods**. Twelve interview-duration values are missing, and no recorded duration exceeds 240 minutes. Recorded respondent BMI values agree with calculations using self-reported weight in kilograms divided by height in metres squared, rounded to one decimal place.

The checks establish internal consistency and agreement with selected questionnaire rules, rather than the accuracy of health and behavioural reports, which were not validated against medical records. Disclosure reduction is separate from these checks. Implications of reporting sources, changing measurement and non-probability recruitment for analysis are described in **Usage Notes**.

### Usage Notes

The RDS files can be opened in R version 3.5.0 or later using ‘readRDS’. For wave-specific analyses, select rows using the ‘wave’ variable and use the corresponding columns, for example, rows with ‘wave = 2’ and variables ending in ‘_w2’. Interpret grouped demographic variables using their category labels, rather than the internal integers R uses to store factors.

Researchers should define their unit of analysis before linking or combining records. Analyses of individual reports should retain the reporting respondent’s identifier and ‘wave’ alongside the alter’s ‘node_code’. Producing one summary per person requires a clear rule for combining reports that may differ. Statistical analyses should account for repeated respondents and shared alters where relevant. Ignoring these dependencies can produce standard errors that are too small and confidence intervals that are too narrow. For longitudinal analyses, derive interview participation from the presence of ‘unique_egoID’ and ‘wave’ in the ego table. An absent interview record is different from a missing item within a completed interview and does not, by itself, establish withdrawal from the study.

Longitudinal comparisons should distinguish fresh elicitation in waves 2 and 3 from historical-roster review by returning respondents in wave 4. A person’s absence from a later list does not necessarily mean that the relationship ended, and a larger reviewed list does not necessarily indicate network growth. Newly added contacts may also represent longstanding relationships. Wave-4 analyses should distinguish first-time from returning respondents and clearly specify any contact-frequency or residence criteria used to select alters.

The alter-alter table was prepared to include only pairs whom the respondent (ego) reported as knowing each other. Each retained row therefore indicates acquaintance according to that respondent. Pairs reported as not knowing each other, or for which the respondent was unsure whether they knew each other, were excluded. Uncertainty about how often two alters interact is different from uncertainty about whether they know each other: records with unknown interaction frequency remain in waves 3 and 4. Wave 4 also retains pairs reported to know each other without interacting. Wave 1 describes friendship or acquaintance, whereas waves 2-4 describe interaction frequency, with different response options. These categories should not be treated as identical measures across waves or compared using their numeric codes alone. Returning respondents did not provide new alter-alter assessments in wave 4, so their missing wave-4 records reflect the collection design.

Missing values can occur because a question was not applicable or was not asked, no answer was provided, a value was removed during data cleaning, or a response was removed to protect personal information. For each analysis, researchers should identify who was eligible to answer the question, report how many responses were available, and explain how missing responses were handled. The data show each person’s age group and income category, not their exact age or income. Changes in income are visible only when a person moves from one category to another. Calculated respondent BMI and ego-assessed photographic alter body shape have different measurement sources and should not be pooled as a single measure.

For recruitment analyses, establish interview participation from the ego table rather than infer it from a referral or alter nomination. The sample does not include all commune residents, and the pooled networks do not capture all relationships in the community. Earlier data deposits were prepared for specific studies and contain selected data or outputs produced for those analyses^**14-19**^. Researchers should cite the version of the dataset used in their analysis. Before combining it with an earlier dataset, they should check that matching codes identify the same people and avoid counting the same interview or network record twice.

#### Ethics statement

The study was reviewed and approved by the Ethics Committee of the Center for Innovation in Medicine (INOMED; decisions D001/09-06-2023, D001/19-01-2024 and D002/03-02-2025). The 2025 decision covered both the March-April and September 2025 fieldwork periods. Participation was voluntary. Written informed consent was obtained from all participants for study participation and for the use and sharing of data for scientific research, including future research.

## Data Availability

The dataset (version 1.0.0) is openly available on Zenodo at https://doi.org/10.5281/zenodo.22935150 under the Creative Commons Attribution 4.0 International (CC BY 4.0) licence. The repository contains five linked data tables in RDS and CSV formats, together with the accompanying documentation described in the Data Records section of the manuscript.

## Data availability

The dataset is openly available at Zenodo (https://zenodo.org/uploads/22935150) under version 1.0.0, licensed under CC-BY-4.0, with DOI: 10.5281/zenodo.22935150. The data deposit contains the five linked tables in RDS and CSV formats, along with the accompanying documentation described in Data Records.

## Code availability

R code for importing Network Canvas interview exports is available in the official Network Canvas documentation (https://documentation.networkcanvas.com/en/analyze-data/working-with-data). After import, the research team manually cleaned the data and assigned person identifiers, as described in Methods. No custom code was used beyond standard data cleaning and descriptive analyses, which are fully described in the Data Descriptor. Accordingly, no standalone software or analysis package has been generated that would require a separate code repository.

## Acknowledgements

We thank Iulian Oană, Andreea-Ştefania Moise, Maria-Alexandra Roşu, Antonin-Tron Lozai for their technical help and contribution to data collection and preparation. Finally, we are grateful to our colleagues from the Center for Innovation in Medicine, the University of Bucharest (Department of Sociology), and the 4P-CAN consortium for their methodological insights and constructive discussions that supported the development of this work.

## Author contributions

**M.-G.H**.: Writing - review & editing, Writing - original draft, Visualisation, Supervision, Software, Methodology, Investigation, Formal analysis, Data curation, Conceptualisation. **M.G**.: Writing - review & editing, Writing - original draft, Validation, Project administration, Methodology, Funding acquisition, Conceptualisation. **J.L.M**.: Writing - review & editing, Writing - original draft, Validation, Methodology, Conceptualisation. **J.L**.: Writing - review & editing, Writing - original draft, Validation, Methodology, Conceptualisation. **M.-C.G**.: Writing - review & editing, Writing - original draft, Visualisation, Investigation, Formal analysis, Data curation. **B.-E.M**.: Writing - review & editing, Writing - original draft, Visualisation, Data curation, Conceptualisation. **B.-A.V**.: Writing - review & editing, Writing - original draft, Visualisation, Data curation. **C.C**.: Writing - review & editing, Writing - original draft, Resources, Project administration.

